# Noninvasive Optical Monitoring of Cerebral Hemodynamics Immediately after Birth in Neonates with Congenital Heart Disease

**DOI:** 10.1101/2024.10.02.24314818

**Authors:** Chloe N. Winston, Madison E. Bowe, Sura Lee, Nicolina R. Ranieri, Anne Ades, Juliana Gebb, Jack Rychik, Maryam Y. Naim, Anna Bostwick, Rodrigo M. Forti, Tiffany S Ko, Elizabeth E. Foglia, Wesley B. Baker, Jennifer M. Lynch

**Affiliations:** Division of Neurology, Children’s Hospital of Philadelphia, Philadelphia, PA 19104, USA; Division of Neonatology, Children’s Hospital of Philadelphia, Philadelphia, PA 19104, USA; School of Biomedical Engineering, Science and Health Systems, Drexel University, Philadelphia, PA 19104, USA; Richard D. Wood Jr. Center for Fetal Diagnosis and Treatment, Children’s Hospital of Philadelphia; Perelman School of Medicine, Philadelphia, PA, 19104; Department of Anesthesiology and Critical Care Medicine, Children’s Hospital of Philadelphia, Philadelphia, PA 19104, USA; Division of Neonatology, Department of Pediatrics, School of Medicine, University of Pennsylvania, Philadelphia, Pennsylvania; Department of Bioengineering, University of Pennsylvania, Philadelphia, PA 19014; Division of Cardiothoracic Anesthesiology, Children’s Hospital of Philadelphia, Philadelphia, PA 19104, USA

**Keywords:** Congenital heart disease, cerebral monitoring, NIRS, optical monitoring, cerebral saturation, cerebral blood flow, neonatal resuscitation

## Abstract

**Objective:** Critical congenital heart disease (CHD) is associated with neuropsychiatric impairment that may stem from altered cerebral hemodynamics. While cerebral hemodynamics are shown to differ in neonates with CHD, the effect of CHD on cerebral physiology earlier in life during the fetal to neonatal transition period is yet to be elucidated. This period of neonatal resuscitation could represent an opportunity for intervention in critical CHD, so we aimed to characterize cerebral physiology immediately after birth in neonates with critical CHD using noninvasive optical monitoring.

**Methods:** This case series analysis included term neonates with hypoplastic left heart syndrome (HLHS; n=2) and with transposition of the great arteries (TGA; n=2) who were born at Children’s Hospital of Philadelphia. Continuous measurements of cerebral hemodynamics were acquired during the first hour after birth with novel non-invasive frequency-domain diffuse optical spectroscopy (FD-DOS) and diffuse correlation spectroscopy (DCS), as well as commercial near-infrared spectroscopy (NIRS). NIRS measures cerebral oxygenation, and the combined FD-DOS/DCS device measures cerebral oxygenation, blood flow, and metabolism.

**Results:** In all four patients, cerebral oxygen saturation was lower than reference values in the literature. Additionally, we observed decreases in cerebral blood flow and oxygen metabolism during postnatal transition that were not reflected by peripheral oxygen saturation. The decreases were spontaneous in infants with HLHS and temporally associated with invasive respiratory support in infants with TGA.

**Conclusion:** This study demonstrates periods of neurological vulnerability during postnatal transition in CHD and motivates further research on the use of noninvasive optical monitoring during neonatal resuscitation.

**Clinical Perspective:** *What is New?:*

- An advanced noninvasive hybrid FD-DOS/DCS system detects periods of neurological vulnerability during neonatal resuscitation in patients with critical CHD that are not reflected by peripheral oxygen saturation trends.
- Invasive respiratory support is associated with cerebral desaturations.

*What Are the Clinical Implications?:*

- These findings suggest the utility of noninvasive cerebral monitoring during neonatal resuscitation in neonates with critical CHD and motivate further studies to assess this.

## Introduction

Children born with critical congenital heart disease (CHD) requiring neonatal cardiac surgery are at an increased risk for poor neurodevelopmental outcomes later in life (1–5). Although this risk is multifactorial, studies have demonstrated that substantial risk exists during the pre-operative period from birth to surgery (6–11). This pre-operative risk may stem from alterations in cerebral physiology in neonates with critical CHD. Previous studies have demonstrated that postnatal cerebral physiology in certain types of critical CHD differs from the physiology in healthy controls (8,12–18). Measurements of decreased fetal cerebral vascular resistance (CVR) in these patients have also been reported (19,20). The cerebral physiology of the fetal to neonatal transition immediately after birth, however, is yet to be elucidated.

Monitoring during neonatal resuscitation immediately after birth is typically limited to arterial oxygen saturation and heart rate, which provides limited insight into cerebral oxygen delivery. To help fill this gap, near-infrared spectroscopy (NIRS) is a promising noninvasive technology for monitoring changes in cerebral tissue oxygenation that has been explored during the transitional period in term and preterm neonates (21–24).

Despite its promise, clinically available continuous-wave NIRS devices are limited in their reproducibility and reliability (25–29). Advanced variants of NIRS technologies are currently being tested for improved quantification. Frequency-domain diffuse optical spectroscopy (FD-DOS) can improve quantification via its direct measurement of tissue scattering properties (30,31). Diffusion correlation spectroscopy (DCS), a qualitatively different near-infrared optical technique, can directly measure cerebral blood flow (32,33). In addition to improved quantification, simultaneous FD-DOS and DCS also provides access to cerebral oxygen metabolism (31,34). Herein, we use a commercial continuous-wave NIRS device and an advanced hybrid FD-DOS/DCS device to monitor transitional changes in cerebral oxygenation, blood flow, and oxygen metabolism in a pilot cohort of neonates with critical CHD. To our knowledge, this case series is the first report of cerebral physiology immediately after birth in neonates with critical CHD.

## Methods

### Study Design

We conducted a case series analysis to evaluate the feasibility of noninvasive cerebral oxygenation and blood flow monitoring using a hybrid FD-DOS/DCS device over the first hour of life in term (>= 37 weeks gestation) neonates with critical congenital heart disease (CHD). This study was conducted in accordance with the ethical principles outlined in the Declaration of Helsinki. The study was approved by the Institutional Review Board (IRB) of Children’s Hospital of Philadelphia, PA. All term neonates who had been prenatally diagnosed with hypoplastic left heart syndrome (HLHS) or transposition of the great arteries (TGA) and who were expected to be born in the Special Delivery Unit (SDU) at Children’s Hospital of Philadelphia were screened for inclusion in the study and parents were approached for consent prenatally. Subjects were excluded if other major congenital anomalies besides CHD were identified, if the neonate was born via stat C-section, if there was multiple gestation pregnancy with concordant sex, if the research equipment could not be set up before birth, if the neonate was delivered to a current COVID-19 positive mother (within 14 days of positive test), or if the neonate was receiving palliative care.

Included patients underwent noninvasive monitoring in the SDU using a commercial continuous-wave NIRS device (Medtronic INVOS 5100C; Minneapolis, Minnesota, USA) and a custom FD-DOS/DCS device. The FD-DOS/DCS instrument and the analysis of its data has been described in detail previously (8,35). The NIRS and FD-DOS/DCS monitoring probes were placed on the patient’s left and right forehead, respectively, as soon as it was feasible with clinical care and within 30 minutes after birth. Regional cerebral tissue oxygen saturation (StO_2_), total hemoglobin concentration (THC), and an index of cerebral blood flow (CBF) were measured using the FD-DOS/DCS device. THC is defined as [*Hb*] + [*HbO*_2_]. Regional cerebral tissue oxygen saturation (rSO_2_) was also measured using the commercial NIRS device. (Note, we use different variables to denote the FD-DOS and NIRS measurements of regional cerebral tissue oxygen saturation to distinguish between the two metrics; in contrast to NIRS, FD-DOS directly measures tissue scattering properties that should in principle improve absolute quantification of tissue oxygen saturation (28,31)). During FD-DOS/DCS and NIRS monitoring, losses of data quality that prevented accurate measurements (*e.g.,* due to large motion artifacts and/or the loss of probe contact) were treated as missing data.

Vital sign data, including heart rate (HR) and preductal peripheral arterial oxygen saturation (SpO_2_) measured with pulse oximetry on the upper right extremity, and rSO_2_ were simultaneously recorded with a bedside vitals acquisition device (CNS-200 Monitor, Moberg Research Inc, Ambler, PA) that was manually synchronized with FD-DOS/DCS measurements. Additional variables evaluated include the cerebral oxygen extraction fraction (OEF) derived from pulse oximetry and FD-DOS data and the cerebral metabolic rate of oxygen (CMRO_2_) derived from FD-DOS, DCS, and blood gas measurement of hemoglobin concentration (Hgb). Specifically, *OEF* = (*SpO*_2_ − *StO*_2_)⁄(γ*StO*_2_), where γ represents the fraction of blood in the venous compartment and is assumed to be 0.75 in this study, and *CMRO*_2_ = 1.39 ⋅ *Hgb* ⋅ *OEF* ⋅ *CBF*_*i*_ where *Hgb* represents the hemoglobin concentration. For the CMRO_2_ measurements, *Hgb* was measured once during neonatal resuscitation and assumed constant for the duration of the monitoring period. OEF and CMRO_2_ were calculated using variables time-averaged across each minute after birth. Dr. Lynch had full access to all the data and takes responsibility for their integrity and the data analysis.

## Results

A total of 34 patients were consented, of which both NIRS and FD-DOS/DCS monitoring were attempted on sixteen patients. Of the sixteen patients, only four neonates had usable FD-DOS/DCS data quality. Data quality in FD-DOS/DCS was limited in the remaining subjects due to loss of probe contact with the skin. Thus, herein we present a case series of four neonates: two with HLHS and two with TGA. These patients were enrolled between July 2021 and March 2023. Subject demographics are summarized in Table 1. All patients were born full-term with a median gestational age at birth of 39.1 weeks (range: 39.0 to 39.3 weeks.) The times after birth for the first acquired FD-DOS, DCS, and NIRS data ranged between 1.3-29.8 minutes, 3.6-18.4 minutes, and 1.3-5.4 minutes, respectively (Table 1).

**Table 1.** Demographic and clinical characteristics of each included neonate. The time of first usable measurement (t_0_) is reported for FD-DOS, DCS, and NIRS data.

| Subject | Prenatal diagnosis | Gestational age at birth (week) | Sex | Delivery mode | FD-DOS $t_0$ (minute) | DCS $t_0$ (minute) | NIRS $t_0$ (minute) |
| --- | --- | --- | --- | --- | --- | --- | --- |
| 1 | HLHS | 39.14 | Female | C-section | 6.8 | 8.2 | 1.3 |
| 2 | HLHS | 39.29 | Male | C-section | 8.9 | 9.4 | 5.4 |
| 3 | TGA | 39.14 | Male | Vaginal | 29.8 | 18.4 | 2.2 |
| 4 | TGA | 39 | Male | Vaginal | 1.3 | 3.6 | 2.7 |

FD-DOS indicates frequency domain diffuse optical spectroscopy; DCS, diffuse correlation spectroscopy; NIRS, near-infrared spectroscopy; HLHS, hypoplastic left heart syndrome; TGA, transposition of the great arteries

Figure 1 shows the continuous multimodal data acquired throughout the first hour of life for the 2 neonates with HLHS (labeled as Subject 1 and Subject 2) who did not receive any inotropic or invasive respiratory support during the first hour of life.

**Figure 1.**
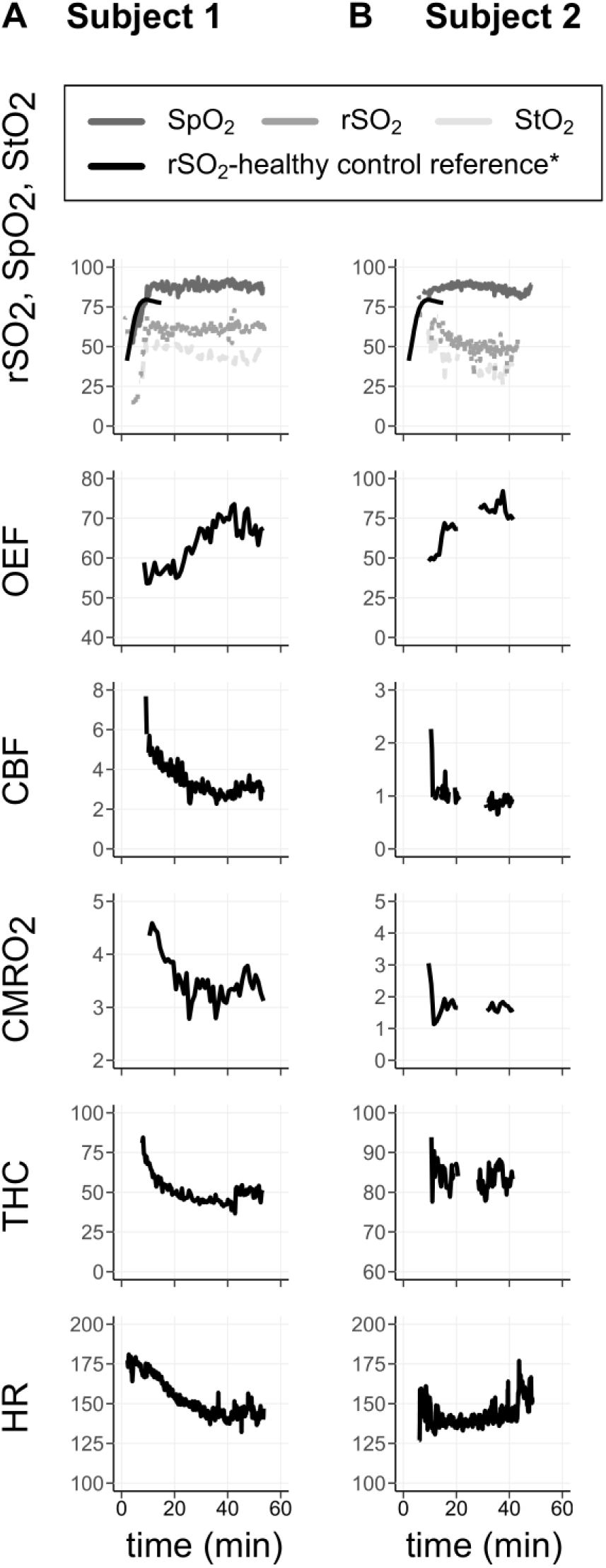
Continuous temporal measurements of cerebral and systemic physiology in two neonates with HLHS during the first hour of life. Continuous measurements include StO_2_, SpO_2_, rSO_2_, OEF, CBF (10^-8^ cm^2^/s), CMRO_2_ (10^7^ml O_2_/dL * cm^2^/s), THC (μmol/L), and HR (beats per minute). The black curves in the oxygen saturation plots are the reference (median) cerebral rSO_2_ measurements after birth previously acquired in a cohort of 381 healthy term neonates requiring no respiratory support during postnatal transition (24).

Systemic and cerebral oxygen saturations increased to peak levels by 15 minutes of life in both neonates. Cerebral oxygen saturations (rSO_2_ and StO_2_) were also noticeably lower than previously reported reference measurements in healthy term neonates requiring no medical support during postnatal transition (24). Interestingly, beginning around 15-20 minutes after birth, an increase in OEF was observed from ∼50% to 75% in both neonates. During this increase in OEF, the SpO_2_ remained stable while CMRO_2_ and CBF decreased. The CBF stabilized at its lower level more rapidly in Subject 2 (OEF also increased more rapidly in this subject.)

Also of interest was the comparison between FD-DOS/DCS-measured StO_2_ and NIRS-measured rSO_2_. In both subjects, StO_2_ was lower than rSO_2_. The saturation trends also differed. In Subject 1, rSO_2_ remained stable after reaching a peak level, but StO_2_ gradually decreased. In Subject 2, both rSO_2_ and StO_2_ decreased, but the decrease was larger in StO_2_. Finally, in Subject 1, THC and heart rate decreased over the monitoring period, whereas in Subject 2, these measurements were more stable.

TGA patients are quite hypoxemic until they receive balloon atrial septostomy and are often sedated during the fetal to neonatal transition period to minimize oxygen consumption. The remaining neonates with TGA (labeled as Subject 3 and Subject 4) underwent invasive respiratory interventions during the monitoring period. Specifically, both subjects were intubated during the first hour of life after premedication with fentanyl and vecuronium. Intubation was initiated with a FiO_2_ of 30% and 100% in Subjects 3 and 4, respectively. Prior to these interventions, Subject 3 was initially put on blow-by oxygen and face mask, and Subject 4 was put on nasal cannula. Subject 3 was then put on mask positive pressure ventilation (PPV) with a fraction of inspired oxygen (FiO_2_) of 50%, and Subject 4 was put on continuous positive airway pressure (CPAP) with FiO_2_ of 100%. The multimodal measurements for these subjects are plotted in Figure 2. As with the HLHS neonates, oxygen saturations increased to peak values over the first 15 minutes of life, and the cerebral oxygen saturations were lower than previously reported reference measurements in healthy neonates requiring no respiratory support (24). In Subject 3, initiation of PPV was followed by a rise in rSO_2_ and a decrease in SpO_2_ and heart rate. FD-DOS/DCS monitoring began after the initiation of CPAP. In Subject 4, rSO_2_ remained fairly stable after initiation of CPAP while SpO_2_ decreased, but FD-DOS/DCS monitoring was limited immediately after initiation.

**Figure 2.**
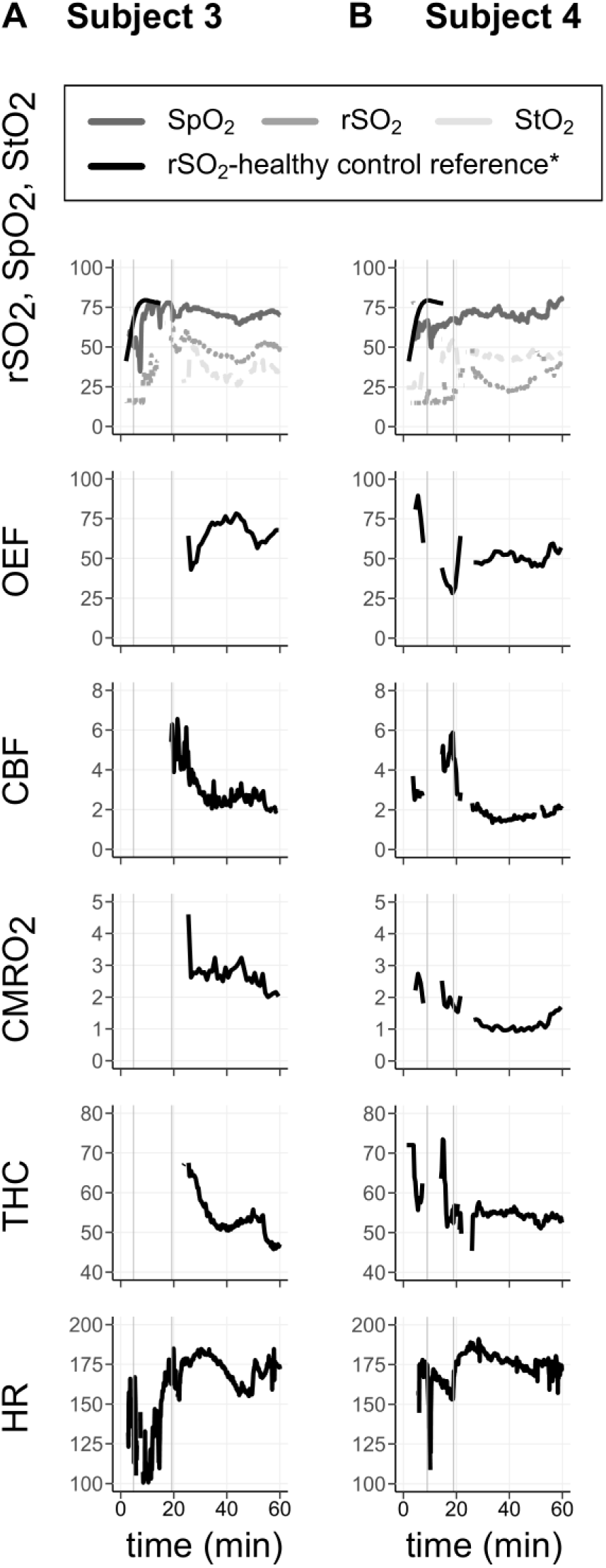
Continuous temporal measurements of cerebral and systemic physiology in two neonates with TGA during the first hour of life. Continuous measurements include StO_2_, SpO_2_, rSO_2_, OEF, CBF (10^-8^ cm^2^/s), CMRO_2_ (10^7^ml O_2_/dL * cm^2^/s), THC (μmol/L), and HR (beats per minute). The black curves in the oxygen saturation plots are the reference (median) cerebral rSO_2_ measurements previously acquired in a cohort of 381 healthy term neonates requiring no medical support (24). Subject 3 and Subject 4 were placed on mask PPV or CPAP, respectively, (first vertical line) and then subsequently intubated (second vertical line).

Interestingly, decreases in CBF were observed following intubations in both subjects. Accompanying changes in cerebral oxygen saturations and total hemoglobin concentration, however, varied. In Subject 3, StO_2_, rSO_2_, and THC all decreased with partial recovery of StO_2_ and rSO_2_ later in the monitoring session. In Subject 4, THC was stable with different trends in the StO_2_ and rSO_2_ data; StO_2_ slightly decreased, while rSO_2_ increased initially with a subsequent large transient decrease. CMRO_2_ and OEF changes following intubation were also variable. In Subject 3, the increased OEF initially compensated for the decreased CBF to maintain CMRO_2_, though the OEF decreased near the end of monitoring and CMRO_2_ decreased. In Subject 4, OEF remained approximately constant, and the CBF decrease was thus matched by an accompanying decrease in CMRO_2_.

## Discussion

Prior longitudinal FD-DOS/DCS monitoring after neonatal transition through the preoperative period in neonates with critical CHD showed impairments in cerebral perfusion and oxygen metabolism (8,14,17,18). However, the impact of critical CHD and its related medical interventions on the fetal to neonatal transition has not been previously studied using noninvasive cerebral monitoring devices. This is the first reported evaluation of cerebral physiology in CHD patients who are only minutes old.

Herein, we used commercial NIRS and advanced FD-DOS/DCS to monitor cerebral blood flow, tissue oxygen saturation, and oxygen metabolism during the first hour of life in four neonates diagnosed with HLHS or TGA. The results of our case series analysis suggest that cerebral physiologic derangements are already present, highlighting the importance of cerebral monitoring during neonatal resuscitation in patients with critical CHD.

During the first 15 minutes of life in all four patients measured, cerebral tissue oxygen saturations were lower than prior reference values obtained in healthy term neonates who received no medical intervention after birth (24), which is consistent with the lower peripheral SpO_2_ of patients with cyanotic CHD (36). Cerebral oxygen saturation trends, however, also provided information that was not apparent in SpO_2_. Specifically, in Subjects 1, 2, and 4, cerebral oxygen saturation decreased while SpO_2_ remained stable. This deviation in trends is notable, as clinicians titrate respiratory support during neonatal resuscitation primarily based on SpO_2_. A stable SpO_2_ trend may thus provide false reassurance to the clinicians on the state of oxygen delivery to and extraction by the brain.

The increases in cerebral oxygen extraction fraction observed in the two patients with HLHS (Subjects 1 and 2) began shortly after measured decreases in cerebral blood flow. Mechanistically, the drop in pulmonary vascular resistance and resultant increase in pulmonary blood flow during the transitional period may cause the reduction in cerebral blood flow, given that HLHS patients have limited antegrade flow. Indeed, the low fetal cerebral vascular resistance of HLHS patients (19,20) suggests they are prone to blood flow reductions because cerebral vascular reserve, (*i.e.,* the capacity to increase flow) is impaired. In both subjects, the increased OEF only partially compensated for the decreased blood flow to sustain metabolism. The observed decreases in cerebral oxygen metabolism, and likely accompanying increases in anaerobic metabolism, suggest increased neurologic vulnerability in these patients.

In the two patients with TGA (Subjects 3 and 4), we observed decreased cerebral blood flow after tracheal intubation. We hypothesize that the increased clearance of CO_2_ after intubation led to cerebral vasoconstriction and decreased cerebral perfusion. The decreased flow corresponded with cerebral desaturation. The heterogeneous responses of peripheral and cerebral physiology to ventilation suggest the utility of cerebral monitoring for guiding ventilation strategies to optimize cerebral blood flow.

There was also heterogeneity across subjects in the agreement between trends in FD-DOS/DCS and commercial NIRS cerebral oxygen saturation measurements. In Subjects 2 and 3, trends between the two oxygen saturation measures were consistent. In Subject 1, however, commercial NIRS rSO_2_ was stable while FD-DOS/DCS StO_2_ decreased, and in Subject 4, a much larger transient decrease in rSO_2_ was observed than in StO_2_. We expect that the FD-DOS/DCS quantification is more accurate because of the technique’s direct measurement of tissue scattering properties (30,31). There is also evidence that commercial NIRS measurements are less accurate at lower tissue oxygen saturations, an observation known as a ‘floor effect’ (25–29). Future work in larger cohorts is needed to compare the utility of FD-DOS/DCS versus commercial NIRS tissue oxygen saturation measurements.

In addition, FD-DOS/DCS measurements were not reliably obtained during neonatal resuscitation in our cohort (data quality was not sufficient in 12 of the 16 patients for which the FD-DOS/DCS sensor was placed). The FD-DOS/DCS sensor is composed of multiple fiberoptic bundles that increase its stiffness and weight relative to commercial NIRS probes. This, in combination with amniotic fluid and vernix on the forehead and significant patient motion, often resulted in the FD-DOS/DCS probe losing contact with the forehead. In future work, improved sensor flexibility and improved cleaning of the forehead will be employed to improve the reliability of FD-DOS/DCS monitoring.

Cerebral tissue oxygen saturation, as well as other hybrid FD-DOS/DCS metrics, (i.e., THC, OEF, CMRO_2_), provide novel insights on cerebral pathophysiology. This case series reports the temporal dynamics of these novel multimodal measurements during the first hour after birth in two neonates with HLHS and two neonates with TGA. Periods of decreased cerebral oxygen saturation and metabolism were observed in the transitional period that were not entirely reflected in peripheral oxygen saturation. These reductions were spontaneous in the HLHS neonates and related to respiratory support in the TGA neonates, possibly resulting from impaired cerebrovascular reserve or autoregulatory capacity. These findings suggest that periods of neurologic vulnerability occur during neonatal resuscitation in patients with critical congenital heart disease and motivate future studies assessing the relevance of cerebral monitoring in this setting.

## Availability of Data and Materials

Data may be made available on reasonable request.

## Author Contributions

CNW: Analysis and interpretation of data, drafting the initial and final manuscript and reviewing it for critical content. MEB: Acquisition, analysis and interpretation of data, reviewing manuscript for critical content. NRR: Software, acquisition of data, validation, reviewing manuscript for critical content. SL, AA, JG, JR, MN, AB: Subject recruitment, echo interpretation to assess eligibility, data management, reviewing manuscript for critical content. RMF: Software, validation, reviewing manuscript for critical content. TSK: Conceptualization, methodology, interpretation of data, software, validation, reviewing manuscript for critical content. EF, WBB, JML: Supervision, conceptualization, methodology, interpretation of data, drafting the initial and final manuscript and reviewing it for critical content.

## Non-standard Abbreviations and Acronyms

CBF: cerebral blood flow

CHD: congenital heart disease

CMRO_2_: cerebral metabolic rate of oxygen

CPAP: continuous positive airway pressure

DCS: diffuse correlation spectroscopy

FD-DOS: frequency domain-diffuse optical spectroscopy

FiO_2_: fraction of inspired oxygen

Hgb: hemoglobin

HLHS: hypoplastic left heart syndrome

IRB: Institutional Review Board

NIRS: near-infrared spectroscopy

OEF: oxygen extraction fraction

PPV: positive pressure ventilation

rSO_2_: regional oxygen saturation

SDU: Special Delivery Unit

SpO_2_: peripheral oxygen saturation

StO_2_: tissue oxygen saturation

TGA: transposition of the great arteries

THC: total hemoglobin concentration

## Data Availability

Data may be made available on reasonable request.

## References

1. Brosig CL, Bear L, Allen S, Simpson P, Zhang L, Frommelt M, Mussatto KA. Neurodevelopmental outcomes at 2 and 4 years in children with congenital heart disease. Congenital Heart Disease. 2018;13:700–705.

2. Butler SC, Sadhwani A, Stopp C, Singer J, Wypij D, Dunbar-Masterson C, Ware J, Newburger JW. Neurodevelopmental assessment of infants with congenital heart disease in the early postoperative period. Congenital Heart Disease. 2019;14:236–245.

3. Kovacs AH, Saidi AS, Kuhl EA, Sears SF, Silversides C, Harrison JL, Ong L, Colman J, Oechslin E, Nolan RP. Depression and anxiety in adult congenital heart disease: Predictors and prevalence. International Journal of Cardiology. 2009;137:158–164.

4. Peyvandi S, Chau V, Guo T, Xu D, Glass HC, Synnes A, Poskitt K, Barkovich AJ, Miller SP, McQuillen PS. Neonatal Brain Injury and Timing of Neurodevelopmental Assessment in Patients With Congenital Heart Disease. Journal of the American College of Cardiology. 2018;71:1986–1996.

5. Schaefer C, Von Rhein M, Knirsch W, Huber R, Natalucci G, Caflisch J, Landolt MA, Latal B. Neurodevelopmental outcome, psychological adjustment, and quality of life in adolescents with congenital heart disease. Develop Med Child Neuro. 2013;55:1143–1149.

6. Licht DJ, Jacobwitz M, Lynch JM, Ko T, Boorady T, Devarajan M, Heye KN, Mensah-Brown K, Newland JJ, Schmidt A, et al. Impaired Maternal-Fetal Environment and Risk for Preoperative Focal White Matter Injury in Neonates With Complex Congenital Heart Disease. JAHA. 2023;12:e025516.

7. Limperopoulos C, Majnemer A, Shevell MI, Rosenblatt B, Rohlicek C, Tchervenkov C. Neurodevelopmental status of newborns and infants with congenital heart defects before and after open heart surgery. The Journal of Pediatrics. 2000;137:638–645.

8. Lynch JM, Ko T, Busch DR, Newland JJ, Winters ME, Mensah-Brown K, Boorady TW, Xiao R, Nicolson SC, Montenegro LM, et al. Preoperative cerebral hemodynamics from birth to surgery in neonates with critical congenital heart disease. The Journal of Thoracic and Cardiovascular Surgery. 2018;156:1657– 1664.

9. Petit CJ, Rome JJ, Wernovsky G, Mason SE, Shera DM, Nicolson SC, Montenegro LM, Tabbutt S, Zimmerman RA, Licht DJ. Preoperative Brain Injury in Transposition of the Great Arteries Is Associated With Oxygenation and Time to Surgery, Not Balloon Atrial Septostomy. Circulation. 2009;119:709–716.

10. Williams IA, Fifer C, Jaeggi E, Levine JC, Michelfelder EC, Szwast AL. The association of fetal cerebrovascular resistance with early neurodevelopment in single ventricle congenital heart disease. American Heart Journal. 2013;165:544–550.e1.

11. Williams IA, Tarullo AR, Grieve PG, Wilpers A, Vignola EF, Myers MM, Fifer WP. Fetal cerebrovascular resistance and neonatal EEG predict 18-month neurodevelopmental outcome in infants with congenital heart disease. Ultrasound in Obstet & Gyne. 2012;40:304–309.

12. Nagaraj UD, Evangelou IE, Donofrio MT, Vezina LG, McCarter R, Du Plessis AJ, Limperopoulos C. Impaired Global and Regional Cerebral Perfusion in Newborns with Complex Congenital Heart Disease. The Journal of Pediatrics. 2015;167:1018–1024.

13. Licht DJ, Wang J, Silvestre DW, Nicolson SC, Montenegro LM, Wernovsky G, Tabbutt S, Durning SM, Shera DM, Gaynor JW, et al. Preoperative cerebral blood flow is diminished in neonates with severe congenital heart defects. The Journal of Thoracic and Cardiovascular Surgery. 2004;128:841–849.

14. Dehaes M, Cheng HH, Buckley EM, Lin P-Y, Ferradal S, Williams K, Vyas R, Hagan K, Wigmore D, McDavitt E, et al. Perioperative cerebral hemodynamics and oxygen metabolism in neonates with single-ventricle physiology. Biomed Opt Express. 2015;6:4749.

15. Charbonneau L, Chowdhury RA, Marandyuk B, Wu R, Poirier N, Miró J, Nuyt A - M., Raboisson M-J., Dehaes M. Fetal cardiac and neonatal cerebral hemodynamics and oxygen metabolism in transposition of the great arteries. Ultrasound in Obstet & Gyne. 2023;61:346–355.

16. Leon RL, Bitar L, Sharma K, Mir IN, Chalak LF. Postnatal Cerebral Hemodynamics and Placental Vascular Malperfusion Lesions in Neonates With Congenital Heart Disease. Pediatric Neurology. 2024;156:72–78.

17. Cheng HH, Ferradal SL, Vyas R, Wigmore D, McDavitt E, Soul JS, Franceschini MA, Newburger JW, Grant PE. Abnormalities in cerebral hemodynamics and changes with surgical intervention in neonates with congenital heart disease. The Journal of Thoracic and Cardiovascular Surgery. 2020;159:2012–2021.

18. Lynch JM, Buckley EM, Schwab PJ, McCarthy AL, Winters ME, Busch DR, Xiao R, Goff DA, Nicolson SC, Montenegro LM, et al. Time to surgery and preoperative cerebral hemodynamics predict postoperative white matter injury in neonates with hypoplastic left heart syndrome. The Journal of Thoracic and Cardiovascular Surgery. 2014;148:2181–2188.

19. Kaltman JR, Di H, Tian Z, Rychik J. Impact of congenital heart disease on cerebrovascular blood flow dynamics in the fetus. Ultrasound in Obstet & Gyne. 2005;25:32–36.

20. Szwast A, Tian Z, McCann M, Soffer D, Rychik J. Comparative analysis of cerebrovascular resistance in fetuses with single-ventricle congenital heart disease. Ultrasound in Obstet & Gyne. 2012;40:62–67.

21. Massirio P, Cardiello V, Andreato C, Caruggi S, Battaglini M, Calandrino A, Polleri G, Mongelli F, Malova M, Minghetti D, et al. Ventilatory Support, Extubation, and Cerebral Perfusion Changes in Pre-Term Neonates: A Near Infrared Spectroscopy Study. Neurotrauma Reports. 2024;5:409–416.

22. Wolfsberger CH, Pichler-Stachl E, Höller N, Mileder LP, Schwaberger B, Avian A, Urlesberger B, Pichler G. Cerebral oxygenation immediately after birth and long-term outcome in preterm neonates—a retrospective analysis. BMC Pediatr. 2023;23:145.

23. Baik N, Urlesberger B, Schwaberger B, Schmölzer GM, Avian A, Pichler G. Cerebral haemorrhage in preterm neonates: does cerebral regional oxygen saturation during the immediate transition matter? Arch Dis Child Fetal Neonatal Ed. 2015;100:F422–F427.

24. Pichler G, Binder C, Avian A, Beckenbach E, Schmölzer GM, Urlesberger B. Reference Ranges for Regional Cerebral Tissue Oxygen Saturation and Fractional Oxygen Extraction in Neonates during Immediate Transition after Birth. The Journal of Pediatrics. 2013;163:1558–1563.

25. Ajayan N, Thakkar K, Lionel KR, Hrishi AP. Limitations of near infrared spectroscopy (NIRS) in neurosurgical setting: our case experience. J Clin Monit Comput. 2019;33:743–746.

26. Barud M, Dabrowski W, Siwicka-Gieroba D, Robba C, Bielacz M, Badenes R. Usefulness of Cerebral Oximetry in TBI by NIRS. JCM. 2021;10:2938.

27. Hamrin TH, Radell PJ, Fläring U, Berner J, Eksborg S. Performance of regional oxygen saturation monitoring by near-infrared spectroscopy (NIRS) in pediatric inter-hospital transports with special reference to air ambulance transports: a methodological study. J Clin Monit Comput. 2018;32:841–847.

28. Lynch JM, Mavroudis CD, Ko TS, Jacobwitz M, Busch DR, Xiao R, Nicolson SC, Montenegro LM, Gaynor JW, Yodh AG, et al. Association of Ongoing Cerebral Oxygen Extraction During Deep Hypothermic Circulatory Arrest With Postoperative Brain Injury. Seminars in Thoracic and Cardiovascular Surgery. 2022;34:1275–1284.

29. McNeill S, Gatenby JC, McElroy S, Engelhardt B. Normal cerebral, renal and abdominal regional oxygen saturations using near-infrared spectroscopy in preterm infants. J Perinatol. 2011;31:51–57.

30. Zhou X, Xia Y, Uchitel J, Collins-Jones L, Yang S, Loureiro R, Cooper RJ, Zhao H. Review of recent advances in frequency-domain near-infrared spectroscopy technologies [Invited]. Biomed Opt Express. 2023;14:3234.

31. Durduran T, Choe R, Baker WB, Yodh AG. Diffuse optics for tissue monitoring and tomography. Rep Prog Phys. 2010;73:076701.

32. Durduran T, Yodh AG. Diffuse correlation spectroscopy for non-invasive, micro-vascular cerebral blood flow measurement. NeuroImage. 2014;85:51–63.

33. Carp SA, Robinson MB, Franceschini MA. Diffuse correlation spectroscopy: current status and future outlook. Neurophoton. 2023;10. doi:10.1117/1.NPh.10.1.013509.

34. Jain V, Buckley EM, Licht DJ, Lynch JM, Schwab PJ, Naim MY, Lavin NA, Nicolson SC, Montenegro LM, Yodh AG, et al. Cerebral Oxygen Metabolism in Neonates with Congenital Heart Disease Quantified by MRI and Optics. J Cereb Blood Flow Metab. 2014;34:380–388.

35. Shaw K, Mavroudis CD, Ko TS, Jahnavi J, Jacobwitz M, Ranieri N, Forti RM, Melchior RW, Baker WB, Yodh AG, et al. The use of novel diffuse optical spectroscopies for improved neuromonitoring during neonatal cardiac surgery requiring antegrade cerebral perfusion. Front Pediatr. 2023;11:1125985.

36. Thomas AR, Ma AL, Weinberg DD, Huber M, Ades A, Rychik J, Foglia EE. Delivery room oxygen physiology and respiratory interventions for newborns with cyanotic congenital heart disease. J Perinatol. 2021;41:2309–2316.

